# UCSF RMaC: University of California San Francisco 3D Multi-Phase Renal Mass CT Dataset with Tumor Segmentations

**DOI:** 10.64898/2026.02.11.26346096

**Authors:** Sule Sahin, Ernesto Diaz, Abhejit Rajagopal, Mansour Abtahi, Samantha Jones, Qing Dai, Sage Kramer, Zhen Jane Wang, Peder EZ Larson

## Abstract

Current standard of care imaging practices cannot reliably differentiate among certain renal tumors such as benign oncocytoma and clear cell renal cell carcinoma (RCC), and between low and high grade RCCs. Previous work has explored using deep learning, radiomics, and texture analysis to predict renal tumor subtypes and differentiate between low and high grade RCCs with mixed success. To further this work, large diverse datasets are needed to improve model performance and provide strong evaluation sets. In this work, a dataset of 831 multi-phase 3D CT exams was curated. Each exam contains up to three contrast-enhanced CT phases. Tumor outlines or bounding boxes were annotated and registered to the image volumes. The pathology results for each tumor and relevant patient metadata are also included.

## Background & Summary

Accurate characterization of localized renal masses at the T1 stage is crucial for proper treatment planning. CT exams are the most common practice for renal tumor diagnosis and staging. These exams usually consist of an unenhanced pre-contrast CT scan, and one or more post-contrast phases with varied delay times after contrast administration. However, even with contrast-enhanced CT, it is difficult to reliably differentiate malignant versus benign renal masses, between the subtypes of renal cell carcinomas (RCCs), and low-versus high-grade RCC^1–4^.

Previously, researchers have explored deep learning^5–11^ and radiomics with texture analysis^12–16^ to differentiate pathologies or tumor grades using unenhanced or contrast-enhanced CT with varying levels of success. These previous studies had datasets smaller than 500 cases and were from a single institution. Inclusion of publicly available datasets from a diverse set of external institutions could improve the results of these techniques and improve the model’s generalization to new data during inference.

Current publicly available multiphase renal mass CT datasets are limited in the number of patient cases. The TCGA-KIRC^17^ (n=267), TCGA-KIRP^18^ (n=33), and TCGA-KICH^19^ (n=15) datasets include clear cell, papillary, and chromophobe RCC CT datasets, respectively, hosted on the Cancer Imaging Archive (TCIA)^20^. The 2023 Kidney and Kidney Tumor Segmentation challenge (KiTS)^21^ hosted by MICCAI also offers a publicly available dataset with a total of 599 cases combined from the 2021 and 2023 challenge (489 training, 110 test) including clear cell RCC, chromophobe RCC, unclassified RCC, angiomyolipoma (AML), and oncocytoma, but only includes a single CT phase, either corticomedullary or nephrogenic. Multiphase datasets may improve algorithm development for classification tasks owing to the varied contrast in the different phase images.

Here, we provide and describe a 3D multiphase renal mass CT dataset with tumor masks including 831 cases gathered from over 15 years of data being reviewed at a single institution.

## Methods

### DICOM Retrieval, Inclusion Criteria & Conversion to NifTI

Between 2002 and 2018, the UCSF pathology database was searched for renal masses ≤7 cm (T1 stage). Exams were included only if the patient had a preoperative CT which included an unenhanced scan and at least one post-contrast scan. All pathology was performed at UCSF. CT scans came from UCSF and from outside institutions in the case subjects were referred to UCSF clinics. The DICOM images were imported from the institution PACS with anonymized and deidentified headers. DICOM data was converted to Neuroimaging Informatics Technology Initiative (NifTI) files for easy volume manipulation. The study was conducted under an Institutional Review Board approved protocol. Informed consent was waived by the IRB as the research involved no more than minimal risk to the subjects and could not practicably be carried out without the waiver.

### CT Parameters

The CT exams retrieved were not limited by protocol, since many of the renal mass diagnoses were from incidental findings. Thus, the studies included are not limited to, for example, a renal mass protocol, but include various abdominal imaging protocols. This resulted in variability in positioning (e.g. supine versus prone), spatial resolution, and post-contrast timings. The average slice thickness was 3.33 mm, with a range from 0.625 to 10 mm. The average in-plane spatial resolution was 0.78 mm, with a range from 0.42 to 1.36 mm.

### Tumor Segmentations & Phase Labeling

A Radiology resident who had 3 years of experience and went on to an Abdominal Imaging Fellowship segmented renal lesions on axial slices and labeled the phase of the post-contrast images as “arterial”, “portal-venous”, or “delay” on the MD.ai annotation tool. They were supervised by an attending Abdominal Radiologist with over 15 years of experience. These labels were chosen because of the variability in CT protocols used (e.g. not all were from a renal mass protocol). The arterial phase included scans timings designated as a corticomedullary phase (30-40 second delay). The portal venous phase included scan timings designated as the nephrographic phase (80-110 second delay). The delay phase included scan timings designated as the excretory phase (3-10 minute delay).

Three types of tumor annotations were used: (1) Polygonal tumor annotation defined by vertices, (2) Bounding box tumor annotation and (3) Bounding box on center slice and center point of tumor on superiormost and inferiormost slice of tumor. Annotation type (2) was used to accelerate the annotation process. Annotation type (3) was used where the tumor boundary was not visible and difficult to annotate, which is common on images without contrast. The final tumor annotations included 6% type (1), 53% type (2) and 41% type (3).

The annotations were exported from MD.ai and converted to NifTI masks. For annotation type (3), the bounding box was extended and linearly interpolated to the superiormost and inferiormost slice using the center markers.

### Registration

Per exam, the post-contrast images and tumor masks were cropped in the slice dimension to match the FOV of the pre-contrast image. Affine registration was performed using the ANTS toolbox to register the post-contrast phase images to the pre-contrast image (affine transform, similarity metric: Mattes Mutual Information). A mutual information similarity metric was used as it has demonstrated superior performance in the medical imaging space. Although deformable transforms (like SyN) were attempted, the affine transform resulted in the best registration in this scenario. The same deformation matrix was applied to tumor masks to register the masks to the pre-contrast image.

Each case was quality controlled to ensure proper registration by visualizing the first, middle and last five slices of the post-contrast images overlaid on the pre-contrast images. In the cases that failed ANTS registration, the Diffusion Imaging in Python (Dipy) package was used instead (affine transform, mutual information similarity metric).

### Final Dataset

The final dataset consists of 831 exams with tumor masks and labels. During the data curation process, 131 exams were excluded in total (**Figure 1**). While our original criteria required at least one post-contrast phase, there are cases with only a pre-contrast scan because of retrieval or registration failures of the post-contrast scan.

**Figure 1:**
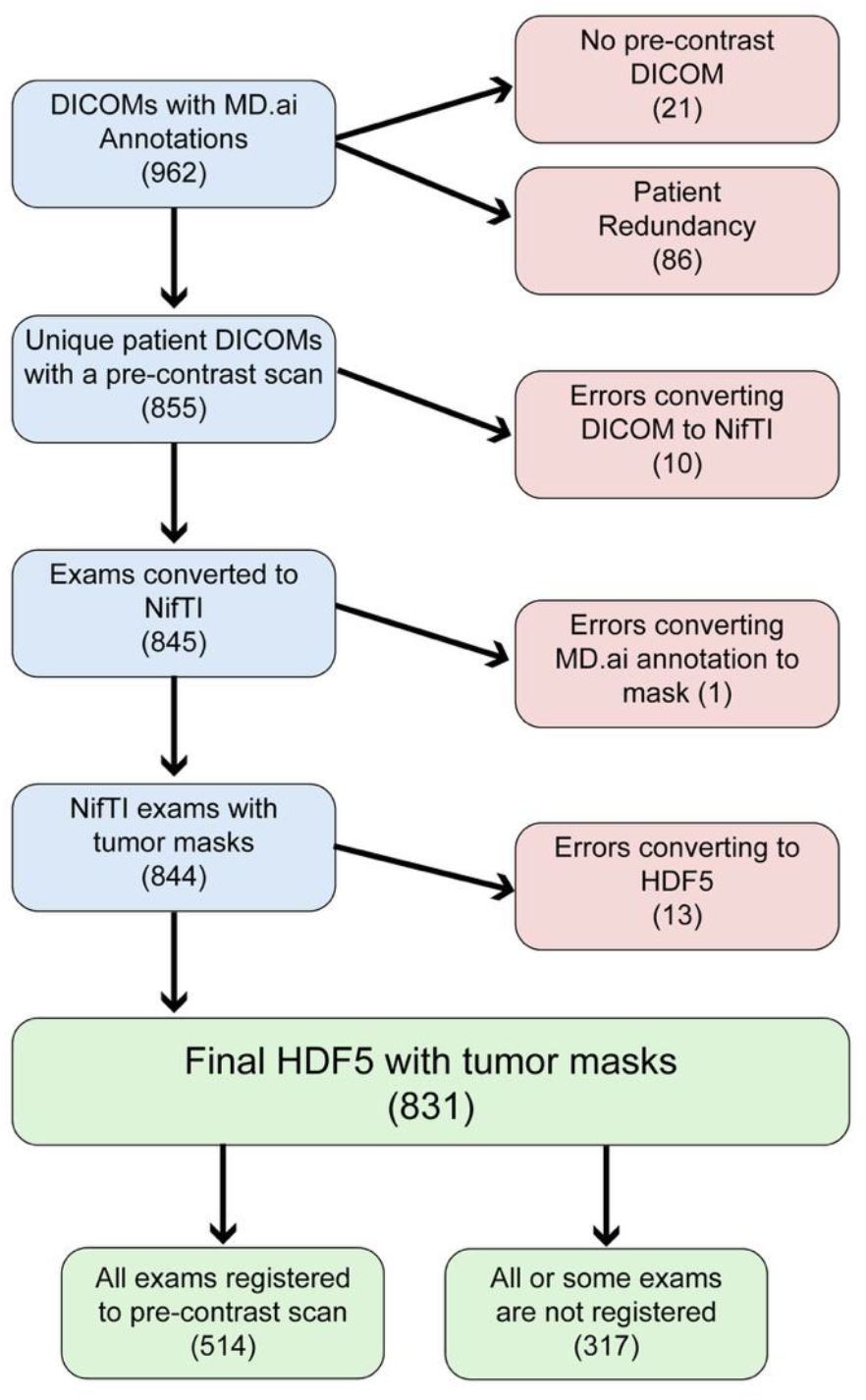
Flowchart of Dataset describing each step of data curation and excluded cases (red boxes). The dataset started with 962 patient DICOMs with annotations and ended with 831 HDF5 datasets that include a tumor mask and tumor labels.

Of the 831 exams, 317 exams failed registration for at least one phase and are included in the dataset without registration. About half the 317 cases that are not registered failed registration because the pre-contrast image was acquired with patient prone on the exam table whereas the post-contrast phase images were acquired with the patient supine on the table, resulting in organ movement and orientation changes. The other cases failed for other reasons, such as imperfect registration, cropped FOV of one or more scans, and failure of registration in the superior/inferior direction.

### Conversion to HDF5

For each exam, the best tumor mask was chosen by annotation type with preference rank in the following order: (1) polygonal segmentation, (2) bounding box on all slices, (3) bounding box on central slice and center markers at end slices. The CT volumes, mask, and relevant metadata (tumor type, pathology, tumor grade) were all saved together as a single Hierarchal Data Format 5 (HDF5) file for each exam. HDF5 files were chosen over NifTI for more metadata flexibility, improved organization and ease-of-use. During the described curation process, each scan was converted from DICOM to NifTI to HDF5. This pipeline could be optimized by directly converting DICOM to HDF5 in the future.

## Data Record

Here, we introduce a dataset consisting of 3D multiphase CT Renal Mass exams. It is freely available through the UCSF Center for Intelligent Imaging (Ci^2^) Datasets for Medical Imaging repository at https://imagingdatasets.ucsf.edu/dataset/3, as well as through the AWS Open Data Registry at https://registry.opendata.aws/ucsf-rmac/. The final dataset contains 831 exams where each exam has a pre-contrast volume, most cases have one or more post-contrast phase volumes (arterial, portal venous and/or delay), a lesion segmentation mask, and pathology metadata (**Figure 2**). Along with CT volumes, masks of the renal masses are also included. Although most of these are bounding boxes, the masks may help localize the tumor quickly for analysis. The CT volumes are provided as-is and no cropping, resampling or normalization has been done to allow for flexibility in preprocessing.

**Figure 2:**
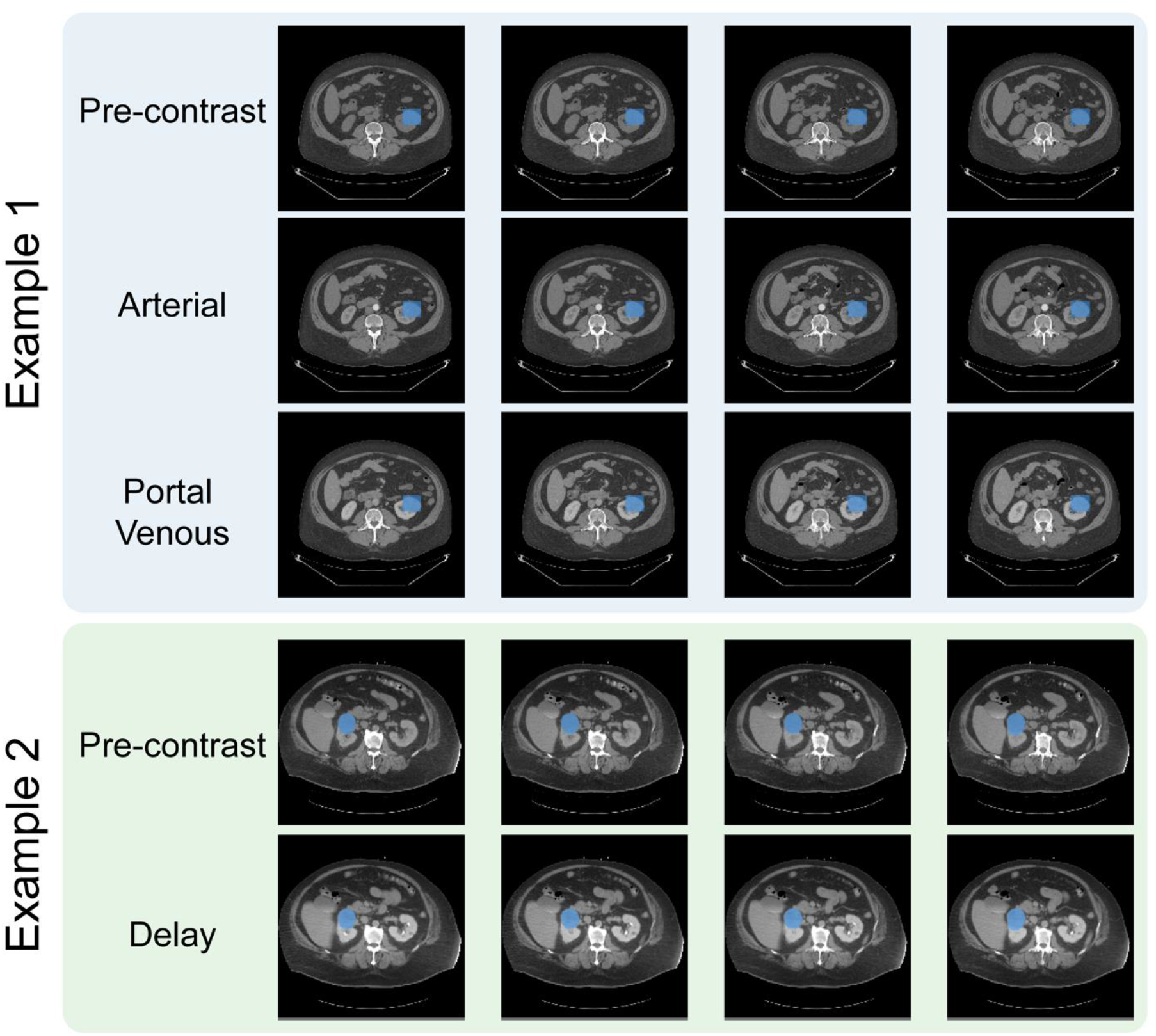
Two example exams from the dataset. For each exam, five slices are shown for multiple phases with the tumor mask shaded in blue. Example 1 shows annotation type (3), a linearly interpolated bounding box. Example 2 shows annotation type (1), a polygonal mask. The dataset stores the volumes without any cropping. The two examples have successful registration.

**Figure 3:**
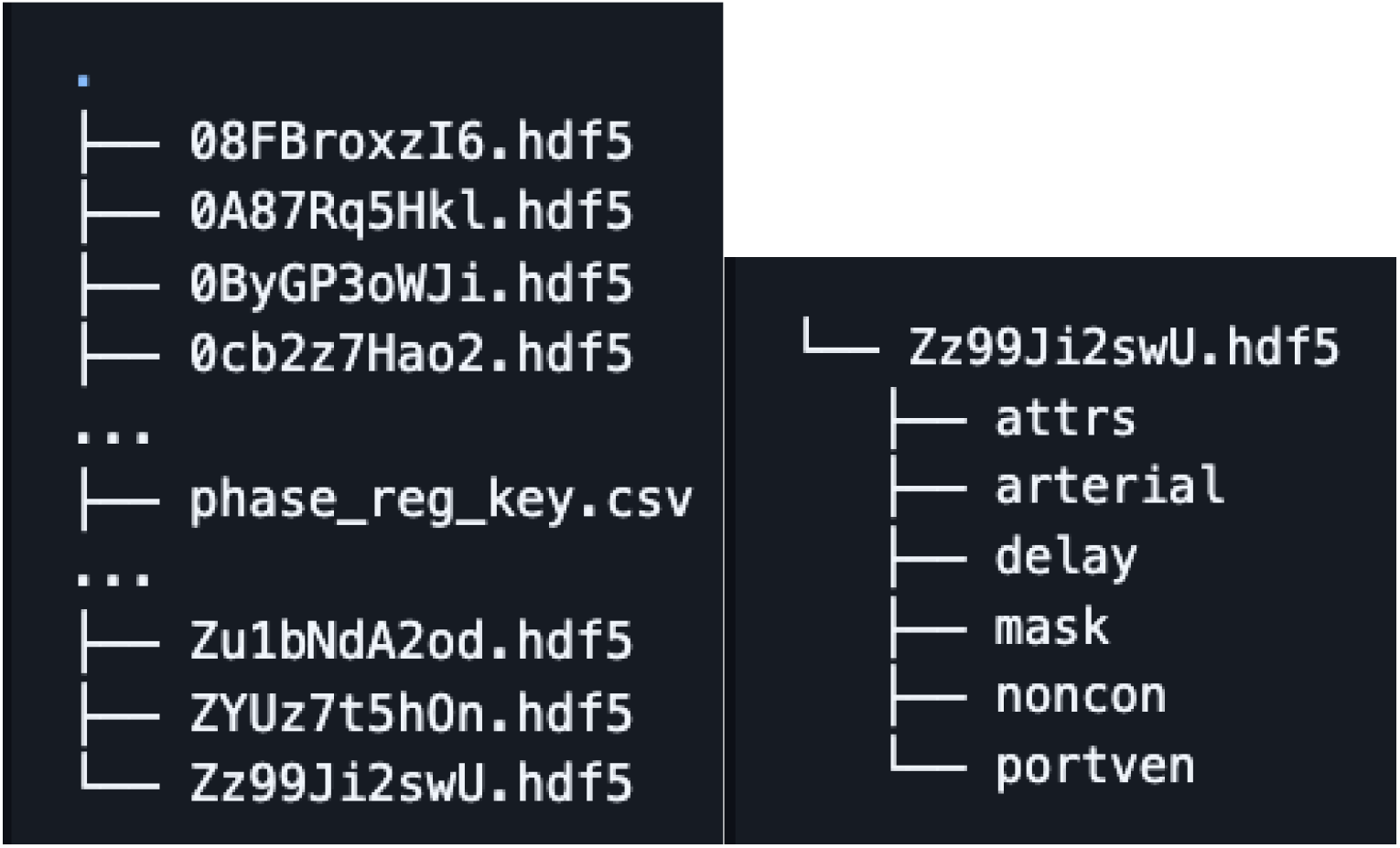
Example file structure of dataset. Dataset includes HDF5 files for each patient and a csv file contained complete metadata. Each HDF5 file contains volumes for the unenhanced CT exam (noncon), any other post contrast exams (arterial, delay, portven), mask of tumor, and attrs containing metadata like patient age, tumor pathology and CT volume pixel dimensions.

The dataset consists of HDF5 files each file containing image data and metadata from a single patient. Additionally, a single csv file is included containing metadata of all the exams. Each exam is indexed with using an anonymized alphanumeric code (referred to as the PID or patient ID).

The csv file includes: registration status of each phase and mask (0=no exam, 1=not registered, 2=registered to unenhanced exam), CT system used, patient age, patient sex, tumor type (RCC, AML, oncocytoma or other), pathology of the tumor, pathology grade (for RCCs), tumor laterality, tumor diameter size in cm and tumor biopsy type . Note noncon= unenhanced CT exam, portven= portal venous exam. For pathology grade, some of the pathologies (like chromophobe RCC) were not given a grade number but labelled more generally as low-(6.0) or high-grade (5.0). Grades 1.0-4.0 represent tumor grade 1-4 respectively.

Each HDF5 file contains the following image volumes unenhanced CT exam volume, 0-3 additional post contrast exam volumes and tumor mask registered to the unenhanced CT exam volume. Each HDF5 files also contains the following metadata as attributes (attrs): CT system used, patient ID, patient age, patient sex, pathology, pathology grade, tumor type and pixel dimensions (*_pixdim) for each CT volume included.

## Technical Validation

To validate the technical quality of the dataset, we performed annotation reviews, analyzed the distribution of the data and annotations, and made measurements of tumor diameter based on the annotations. These results were compared to prior studies. We also provide code for reproducing our data curation process and for reviewing the data to support independent technical validation and review.

All annotations were reviewed by attending Abdominal Radiologist with over 15 years of experience prior to their inclusion in the dataset.

The patient population the dataset was gathered from had an average age of 59.2 years (standard dev: 13.3 years, min: 19 years, max: 93 years). The patient population was majority male (61.3%, female: 38.7%). These are consistent with a recent population-based study that should the highest incidence of renal cancer amongst those aged 55-69 (40.59%), and the majority of cases were men (62.95%)^22^. The laterality was 51.4% right, 45.6% left, and 3.0% bilateral.

The dataset includes a variety of renal mass pathologies, but most are RCCs (**Figure 4**). The most common pathologies were clear-cell RCC (ccRCC, 60.0%), oncocytoma (12.4%), papillary RCC (11.1%), chromophobe RCC (4.8%), and angiomyolipoma (AML, 3.5%). This distribution is consistent with other published work, including a review of resected renal masses showed rates of ccRCC of 68.3%^23^, with another study showing benign mass rates of oncocytoma of 12.3% and AML of 4.4%^24^.

**Figure 4:**
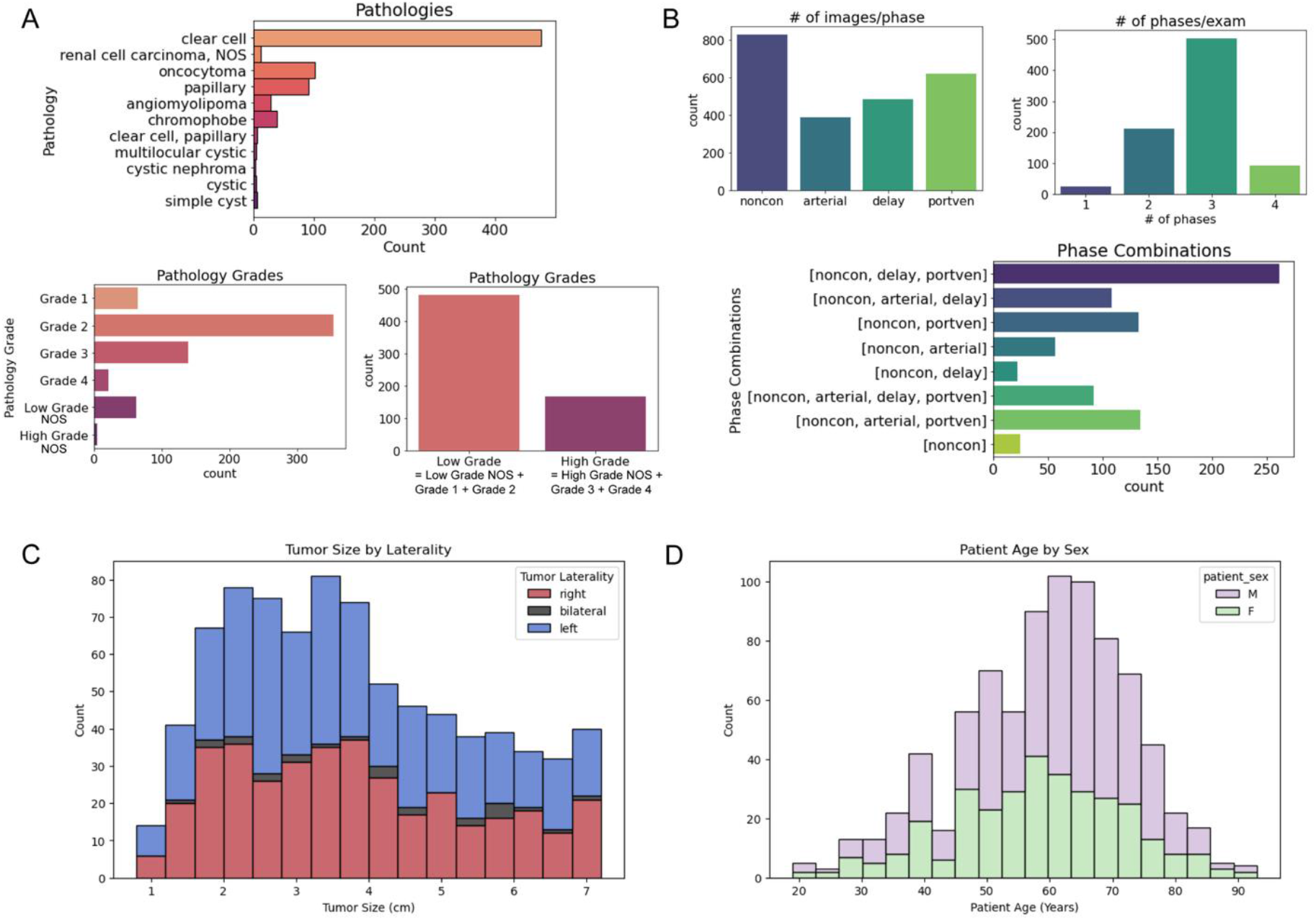
(A) The pathologies & tumor grade labels of the final dataset. Some of the pathologies (like chRCC) were not given a grade number but labelled more generally as low- or high-grade. The majority of the dataset was made up of clear-cell RCC and many of the graded cases were low grade with very few cases being of the highest grade, grade 4. (B) The number of phases and phase combinations of the final dataset. Most exams included the pre-contrast scan and one or two post-contrast phase scans. The most common post-contrast phase was portal venous, although the number of images was similar for all three post-contrast phases. (C) Distribution of tumor diameter size across the dataset, (D) Distribution of patient age. NOS= not otherwise specified, noncon = pre-contrast, portven = portal venous

The pathology grade distribution of RCCs were distributed as grade 1 10.0%, grade 2 54.6%, grade 3 21.5%, and grade 4 3.4%. Similar studies in the literature have reported distributions of grade 1 5.6%, grade 2 66.8%, grade 3 14.1%, and grade 4 1.3%^25^. These similarities further validate the relevance of this dataset.

The tumor segmentations were used to measure the mean tumor diameter to be 3.8 cm in the dataset, with the distribution shown in **Figure 4c**. Note that the dataset only includes renal masses ≤7 cm (T1 stage). This diameter is consistent with cancer registry based measurements for 22,204 T1 stage masses, where the mean tumor size was 4.1 cm^26^.

The data curation code is provided for independent technical validation. Also provided are tutorials describing the annotations and tools for navigating the data are also available in support of independent technical validation and reproducibility, and these can be used to replicate the data distribution analyses and annotation overlay figures shown. They are available open-source at https://github.com/LarsonLab/UCSF-RMaC.

## Data Availability

All data produced are available online at UCSF Center for Intelligent Imaging (Ci2) Datasets for Medical Imaging repository as well as the AWS Open Data Registry.

https://imagingdatasets.ucsf.edu/dataset/3

https://registry.opendata.aws/ucsf-rmac/

https://github.com/LarsonLab/UCSF-RMaC

## Usage Notes

Registration was a significant bottleneck in the data curation and about 30% of exams failed registration. Particularly, the cases where the pre-contrast image was acquired with the patient prone on the table, and the contrast-enhanced images were acquired with the patient supine, proved to be a challenge. In this work, a select number of registration tools were attempted, including both affine and deformable registrations. However, registration tools fine-tuned on abdominal CT application were not attempted which could yield improved results for the remaining exams that require registration. The dataset would benefit from inclusion of additional segmentations from additional readers in order to provide enhanced confidence and allow for assessment of inter-observer agreement. For the addition of new data, the functions and scripts used for the described curation steps are available on GitHub.

## Data Availability

Data generated by the authors or analyzed during the study are available at the UCSF Center for Intelligent Imaging (Ci^2^) Datasets for Medical Imaging repository at https://imagingdatasets.ucsf.edu/dataset/3, as well as through the AWS Open Data Registry at https://registry.opendata.aws/ucsf-rmac/.

## Code Availability

Code is available at the following link https://github.com/LarsonLab/UCSF-RMaC and is shared publicly under a MIT license. The repository contains jupyter notebooks and python scripts used for data curation and tutorials on getting started with using the data. Python 3.6 was used when initially developing the code but more detail about software versions is logged in the environment.yml file.

---

**Supplementary Figure 1:**
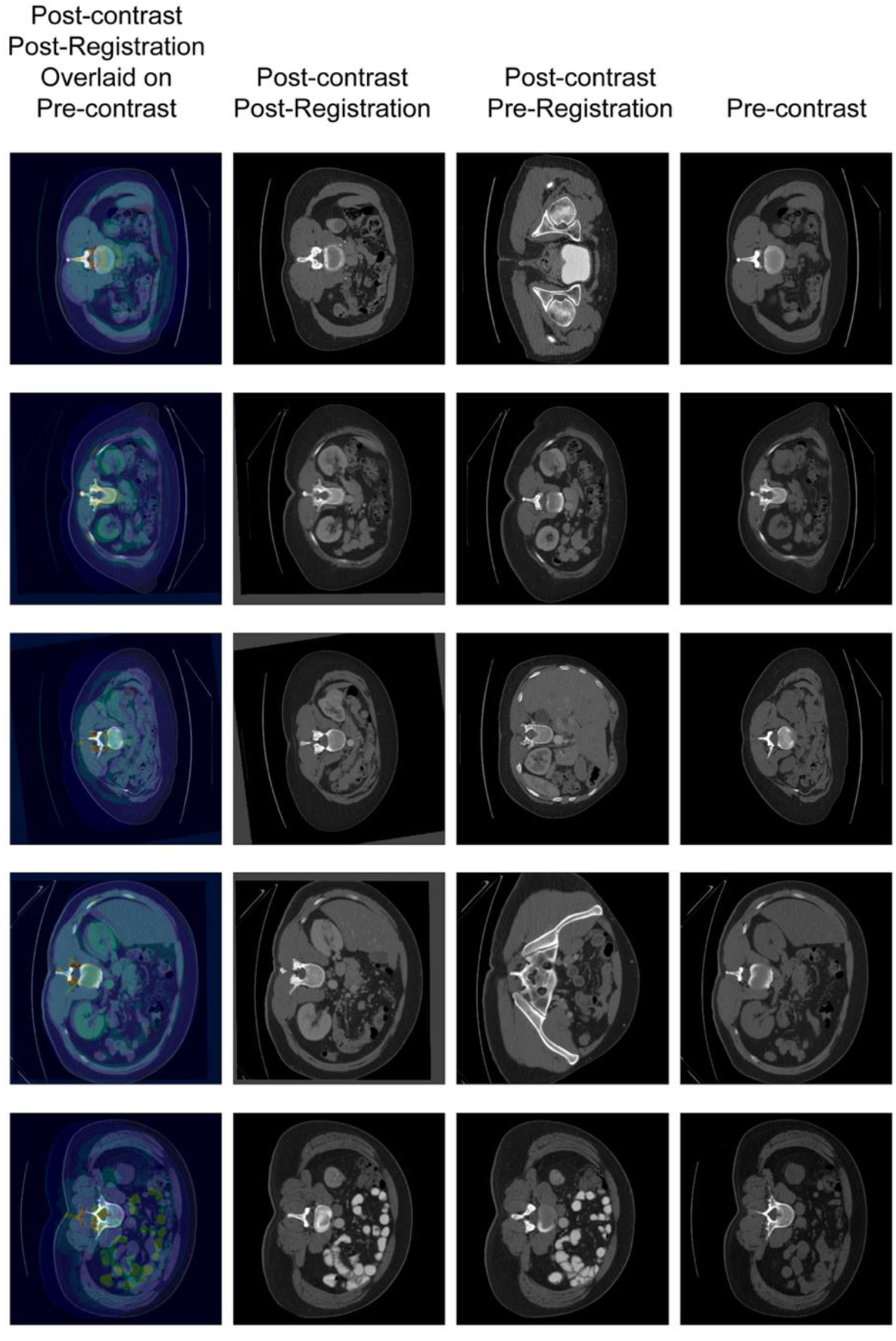
Example of a failed registration. The pre-contrast scan was taken with the patient positioned prone on the CT table and the post-contrast scan here was taken with the patient supine on the table.

